# How to predict relapse in leukemia using time series data: A comparative in silico study

**DOI:** 10.1101/2020.12.04.20243907

**Authors:** Helene Hoffmann, Christoph Baldow, Thomas Zerjatke, Andrea Gottschalk, Sebastian Wagner, Elena Karg, Sebastian Niehaus, Ingo Roeder, Ingmar Glauche, Nico Scherf

**Affiliations:** Institute for Medical Informatics and Biometry, Carl Gustav Carus Faculty of Medicine, School of Medicine, TU Dresden, Fetscherstr. 74, 01307 Dresden, Germany; AICURA medical GmbH, Bessemerstr. 22, 12103 Berlin, Germany; National Center of Tumor Diseases (NCT), Partner Site Dresden, 01307 Dresden, Germany; Max Planck Institute for Human Cognitive and Brain Sciences, Stephanstr. 1a, 04103 Leipzig, Germany

**Author notes:** These authors contributed equally (Helene Hoffmann, Christoph Baldow).

## Abstract

Risk stratification and treatment decisions for leukemia patients are regularly based on clinical markers determined at diagnosis, while measurements on system dynamics are often neglected. However, there is increasing evidence that linking quantitative time-course information to disease outcomes can improve the predictions for patient-specific treatment responses.

We designed a synthetic experiment to compare different computational methods with respect to their ability to accurately predict relapse for chronic and acute myeloid leukemia treatment. Technically, we used clinical reference data to first fit a model and then generate de novo model simulations of individual patients’ time courses for which we can systematically tune data quality (i.e. measurement error) and quantity (i.e. number of measurements). Based hereon, we compared the prediction accuracy of three different computational methods, namely mechanistic models, generalized linear models, and deep neural networks that have been fitted to the reference data.

Our results show that data quality has a higher impact on prediction accuracy than the specific choice of the particular method. We further show that adapted treatment and measurement schemes can considerably improve the prediction accuracy.

Our proof-of-principle study highlights how computational methods and optimized data acquisition strategies can improve risk assessment and treatment of leukemia patients.

## Introduction

Myeloid leukemias are characterized by aberrations affecting the proliferation and maturation of myeloid progenitor cells, leading to the progressive displacement of functional blood cells by immature and dysfunctional *leukemic* cells. Depending on the time scale of the displacement process, myeloid leukemias are further divided in chronic and acute leukemias. The incidence for both subtypes increases strongly with age [1,2].

Patients with chronic myeloid leukemia (CML) typically carry a disease-specific chromosomal translocation forming the *BCR-ABL1* fusion gene [3–6]. Tyrosine kinase inhibitors (TKI) have been established as a targeted therapy leading to molecular remission in most patients under continuous drug administration [7] and substantially raised the 5-years survival rates to above 90% [8]. Molecular monitoring of disease-specific *BCR-ABL1* mRNA in peripheral blood is the established strategy to quantify the leukemic burden under ongoing therapy. Current therapeutic challenges include the cessation of TKI treatment, upon which about 50 % of CML patients develop a molecular recurrence and do not maintain treatment-free remission [9–11].

Acute myeloid leukemia (AML) is a highly heterogeneous disease with a variety of mutational profiles involved [12]. Commonly, a cyclic induction therapy with cytotoxic drugs such as cytarabine and anthracyclines aims to achieve sustainable remission, while a subsequent consolidation therapy supports the maintenance of the remission status. Molecular detection of mutated oncogenes or their transcripts is increasingly used to monitor leukemic burden in treated AML patients and can help to prospectively identify patients at the onset of disease recurrence [13,14].

Disease recurrence after treatment-induced remission is a significant risk for all leukemia patients. Although the reappearance of CML after TKI cessation can be targeted well by restarting the treatment, physical and psychological side effects of retreatment can be minimized if a prospective identification of ineligible patients could be achieved. AML relapse usually occurs after completion of intensive chemotherapy treatment [15] and is associated with a poor prognosis [16]. In those case, the ability to prospectively predict the risk and timing of relapse or molecular recurrence is of highest importance to optimize and adjust the individual treatment strategy.

Currently, treatment decisions are based on the recommended risk stratification schemes. Those risk assessments are commonly based on *static* measurements from single time points, often at diagnosis [17,18]. In contrast, treatment response dynamics, such as the speed of initial remission, are only rarely evaluated for risk stratification [19]. However, it was shown that molecular disease dynamics indeed correlate with therapy response and future relapse occurrence [20–25]. We reason that the direct integration of molecular response dynamics in the form of time-series data, which are increasingly available from standard disease monitoring, is a crucial element to improve the patient-specific risk stratification.

Assessing this question from a technical point of view, there are several, conceptually different approaches to integrate time-series data from molecular disease monitoring into an improved risk assessment. It is so far not clear how well these approaches are suited for time course data of hematological malignancies, and what their particular strengths and weaknesses are in this context. In order to address this question, we study three methods representing typical examples of the methodological spectrum:

- *Mechanistic models* (MM) describe the molecular disease dynamics as a functional consequence resulting from the interaction between relevant system components (such as cell types, drugs, cytokines etc.). They are commonly implemented as systems of ordinary differential equations (ODE) or as stochastic models. While some model parameters might be directly measurable, other model-specific parameters are obtained by optimally fitting the simulated time course to the available patient data. Evolving the model further in time allows to simulate the expected future behavior. Although MMs require considerable expert knowledge about the underlying mechanisms, the results of these models are readily interpretable as the model parameters typically carry explicit biological meaning.
- On the other end of the spectrum, deep learning approaches [26–28] use generic *neural network models* (NN) to adapt them on a training data set for which time-series data and the corresponding future behavior is known. Roughly speaking, the NN implicitly identifies characteristic features within the time course data that correlate with future outcomes. Those methods require no *a priori* knowledge about the underlying mechanisms, but they are not suitable to directly interpret underlying biological mechanisms. Moreover, the training of NN requires a sufficient amount of annotated data.
- Classical statistical models like logistic regression classifiers can be used to correlate characteristic, predefined features of the time course data (such as speed of remission or remission level) with the known outcome. Such statistical models are summarized as *generalized linear models* (GLM) [29]. Herein, prior knowledge about general treatment dynamics is directly incorporated as an explicit feature of the GLM, while no understanding of the underlying biological mechanisms is required. Although GLMs are typically easier to interpret than neural networks (as the influence of parameters on the prediction can be assessed [30]) this probabilistic approach does not allow for explicit mechanistic interpretations as it is the case for MMs.

In this work, we systematically compare these three methods. In particular, we study the influence of data size, sampling density and measurement error on their prediction accuracy. As available data sets of relevant molecular time courses for AML and CML are currently limited, we first generate an artificial patient cohort (*synthetic data*) using established mathematical models of those diseases [23,31] (Fig.1). This artificial data set closely mimics the features of a smaller sample of real patient time courses, while the number of measurements and the particular noise level can be varied systematically and consistently. Based on this reference simulations, we are further able to suggest alternative disease surveillance schemes that may enhance the predictive power.

**Fig. 1:**
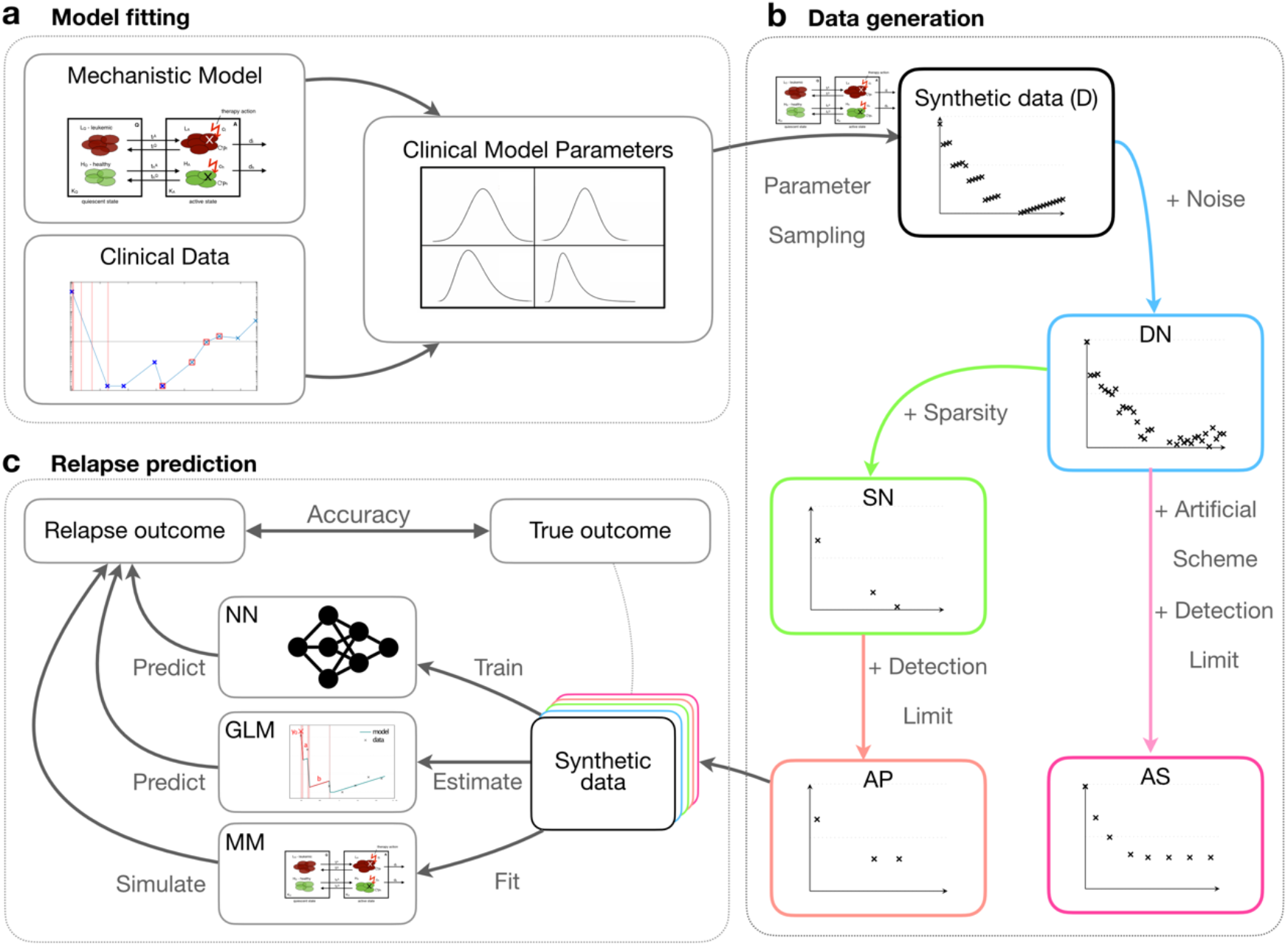
Conceptual overview of our methodological approach: (a) We developed mathematical models for both AML and CML from mechanistic and empirical knowledge [23,31]. The models are first fitted to actual patient data to obtain realistic parameter distributions. (b) We sampled from these empirical parameter distributions to simulate dense, synthetic data (D). We gradually reduced the data quality to mimic actual clinical measurements by introducing noise (dense-noisy, DN), introduce sparsity (sparse-noisy, SN) and a minimum detection limit (artificial patient data, AP). Additionally, we introduced a more informative scheme (artificial scheme, AS), in which the temporal measurements are optimally spaced (AML) or a period of reduced treatment dose precedes therapy cessation (CML). (c) We systematically compared the performance of our mechanistic model (MM), a generalized linear model (GLM) and a neural network (NN) to predict the outcome (relapse/no relapse) of our virtual patient data with varying quality.

## Materials and Methods

### Mechanistic models

To generate the synthetic data, we used our recently published mechanistic models for AML [23] and CML [31], both implemented as systems of ordinary differential equations (ODE). For the AML scenario, four ODEs are used to describe both leukemic and healthy stem cells. Two out of 11 model parameters are optimized to account for patient-specific differences in the disease characteristics, while the others were chosen to account for the general treatment dynamics. For the CML models, three ODEs represent active and inactive leukemic cells plus a population of interacting immune cells. In this case, we estimate 7 of 13 model parameters to optimally describe a patient’s response. Details of the model setup are provided in the Supplementary Text, Suppl. Fig. 6, 7.

### Patient data

For the generation of a set of realistic parameters, we fitted the respective mechanistic model to previously published time course data reflecting the patient’s leukemia remission during and after therapy. In particular, we used the time courses of 275 NPM1-mut AML patients, in which the level of NPM1-mut/ABL abundance is used as a measure of leukemia load (median follow-up time of 10 months, the median number of 5 measurements [23]). Furthermore, we integrated data sets from 21 CML patients reflecting both their BCR-ABL1/ABL1 remission levels under TKI therapy and after therapy cessation (median follow-up time of 84 months, the median number of 28 measurements [31]). Examples of model fits to patient data, and the mean absolute error for each fitted patient can be found in Suppl. Fig. 1.

### Parameter fitting

Both, the AML and the CML model are initially fitted to the available patient data. Technically, we vary possible configurations of free parameters of the model such that the difference to the data is minimized (measured in terms of the sum of squares of the residuals on the logarithmic axis). While a simple optimization routine (*sequential quadratic programming*) is sufficient for the AML models, we apply a genetic algorithm combined with a gradient-based method for the CML scenarios which is better suited to avoid local minima. For further details we refer to the Supplementary Text.

The same optimization routines are applied when the MMs are fitted to the artificial reference data for which we can tune data density and measurement noise (see below).

### Generation of artificial data

To generate artificial patient data, we take random samples from the sets of parameters that were initially derived from fitting the mechanistic models to the available patient data.

In the case of AML, it was sufficient to randomly sample new parameter combinations from the set of empirically observed parameters plus adding a small, normally distributed variation to prevent the generation of identical duplets (see Supplementary Text). For the treatment regime (namely the number and timing of induction cycles) we sampled one particular clinical chemotherapy schedule which we observed in the given patient data. Only artificial patients that reached remission (i.e. leukemic burden fell below the threshold of 1%) were included in the data sets. Using this parameterization and the corresponding schedules, we simulated artificial time courses of 24 months length. In analogy to the clinical situation, AML relapse is assigned if the fraction of leukemic increases above the threshold of 1% within 2 years after treatment start.

For the corresponding artificial CML time-courses, we sampled the seven model parameters from the distribution of empirical estimates in the available data basis under the condition that their mutual correlations are maintained (for details see Supplementary Text). The time of therapy cessation was sampled based on kernel density estimates from the cessation time of the given patients (avg of 92 months with a standard deviation of 28.2 months). This information was then used in de novo forward simulations to generate artificial time-courses of varying duration until treatment stop plus 10 years thereafter. CML recurrence was defined as leukemia abundance > 0.1% (corresponding to BCR-ABL1/ABL1 = 0.1%, MR3).

In order to study how the data quality influences the prediction quality, we generated the following five reference data sets for both disease scenarios (examples in Suppl. Fig. 2 and 3):

- Dense data (D): with weekly (AML) or monthly (CML) exact measurements, respectively.
- Dense-noisy data (DN): where white noise was added to each measurement, according to the noise level found in the given clinical patient data.
- Sparse-noisy data (SN): generated from the DN data set by reducing the number of data points to reflect the measurement frequency in clinical patient data.
- Artificial-Patient data (AP): by adding a detection limit to the SN data as found in the clinical patient data.
- Artificial scheme data (AS): Similar to AP data but using an improved sampling scheme compared to the clinical patient data. For AML measurements are made at the end of each chemotherapy cycle and every six weeks afterwards. For CML, the treatment dose is reduced to half of the usual dose 12 months before therapy cessation with frequent measurements during this period.

Using this synthetic reference data, we use the following setup to evaluate the correctness of predictions. For AML, all measurements from the initial treatment phase to 9 months after diagnosis are provided to the three methods and a corresponding relapse prediction within the subsequent 15 months is derived. For CML, we use all measurements up to the treatment stop to predict whether a patient will present with disease recurrence within ten years thereafter. The long timespan has been chosen to reflect the slow evolution of CML. To obtain the corresponding model predictions from the MM, we fitted the model parameters to the initial time course data (see above) and then simulated the future behavior using the fitted model parameters for each dataset individually. In contrast, both GLM and NN are optimized using a 10-fold cross validation on labelled data sets for which the respective outcome of relapse occurrence is provided.

### Explicit features of time series for GLM analysis

As the Generalized Linear Model, we use a logistic regression classifier. The model uses explicit features that describe characteristics of the time-course data. We took the two characteristics of AML time-courses defined in our previous work [22]: the elimination slope α, describing the speed of decrease of leukemic burden over the time of treatment and the lowest measured leukemic burden after treatment *n*. In this work, we further added three additional features obtained from a segmented regression approach: the leukemic burden at diagnosis (*y*_*0*_, the following decreasing slope during the times of treatment (*a*) and the increasing slope of the leukemic burden in between treatment cycles (*b*) (Suppl. Fig. 4A).

For CML, we defined seven features from fits of a bi-exponential function that described the decrease of the leukemic burden after treatment start. These features include the bi-exponential parameters (*A, α, B, β*), the corresponding deviation of the fit and the data (σ), the cessation time and the BCR-ABL1 value before cessation or half dose. For the AS data, we expand these features with the behavior of the leukemic burden during the time of dose reduction including linear function parameter (*γ*), the deviation during half dose (*C*) and the last measured value before cessation (Suppl. Fig. 4B).

### Neural Network

NN were only trained on the raw time course data with no explicit features provided. To predict the occurrence of relapse, we used a bidirectional Long-short-term-memory (LSTM) network as a default architecture to handle sequence data with varying length. The model consists of a bidirectional LSTM layer followed by a fully connected feature extractor and a binary classification output. We use the respective cross-entropy loss to train the network. We implemented the network in Python using the Keras library [32]. To get a robust estimate of the model performance, we conducted 10 training runs on the same dataset and chose the network with the highest validation accuracy. We then did 10-fold cross-validation for the entire experiment to assess the average and the variability of the results. Further details about the network architecture and training can be found in the Supplementary Text.

### Accuracy

We use the traditional definition of accuracy as the ratio of the number of correct predictions over the total number of predictions: 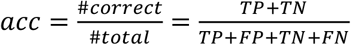 where TP, TN, FP, and FN are true positives, true negatives, false positives and false negatives respectively.

## Results

### Artificial patient data provide a suitable basis to systematically analyze the performance of predictive, computational models

We apply two mechanistic, mathematical models to simulate the dynamics of AML and CML [23,31] thereby creating sets of artificial response data. To make sure that the artificial data resemble real patient time-courses as closely as possible, we fitted the models to respective data sets obtained from 275 AML patients carrying a traceable NPM1-mutation (consisting of a total of 1567 measurements quantifying the relative amount of NMP1-mut transcript [23] over time on a log10-scale) and 21 CML patients (with in total 478 measurements [31] quantifying the relative amount of BCR-ABL transcripts over time on a log10-scale). We report on the overall fitting quality in Suppl. Fig. 1. The fitted model parameters are used to simulate synthetic time courses (Fig. 1a,b). To assess the influence of data quality, we gradually degraded the fully sampled, noise-free time series. We used estimates of the measurement frequencies and measurement errors obtained from the patient data to adjust the corresponding sampling density and noise level for the synthetic data (see Supplementary Text). In total, we created four different datasets with 5000 time-courses from each model to systematically study the influence of data quantity and quality: (i) a dense (D) data set consisting of weekly (AML) or monthly (CML) measurements of the leukemic burden free of any measurement error. (ii) For the dense-noisy (DN) data we added a normally distributed “technical” noise (see Supplementary Text) to all data points of D to match the measuring error (AML) or the residuals observed between real data and their corresponding model fits (CML). (iii) In a third step, we reduced the total number of measurements per patient, creating a sparse-noisy (SN) data set that matches the measurement frequency in the real data. (iv) Finally, to make the data as realistic as possible, we also added a detection limit for very low measurements, called artificial patient (AP) data. Example time courses for all data sets can be found in Suppl. Fig. 2 and 3.

To verify that the created artificial patient data (AP) sets are indeed similar to the real patient data, we derived characteristic features to quantitatively compare them. Those characteristic features refer to typical time scales and remission levels of the patient’s response (see Suppl. Fig. 4, Materials and Methods). The features are computed separately for the AP data and the given patient data. The visual comparison in Suppl. Fig. 5 indicates that the median values of the characteristic features are very similar between AP and real data. It appears, that especially for the case of CML, the synthetic data sets yield a larger variance compared to the real data. A closer look at the data reveals that this is effect, at least partially, results from a sampling effect, as the variance measurement is only based on a small data set (n=21) of real patients.

### Data quality has a strong influence on prediction accuracies, but the drop in performance considerably differs between models and use-cases

Similar to the clinical presentation, we classified the synthetic time-courses as whether they show a relapse or not. For both CML and AML, we define disease recurrence by an increase of the leukemic burden (measured in terms of relative transcript abundance) within a predefined period above a given threshold (AML: leukemic burden increasing > 1% after treatment termination; CML: leukemic burden > 0.1% for at least one month).

We then systematically compared the accuracy of relapse predictions between the three general methods (namely MM, GLM, NN). To do so we provide each method with data from the initial treatment phase and compare the resulting predictions with the ground truth from the artificial data sets. For AML, we provide all measurements from the initial treatment phase until 9 months after diagnosis and derive a corresponding prediction on whether a relapse is expected within the subsequent 15 months. For CML, we use all measurements up to treatment stop to predict whether a patient will present with disease recurrence within ten years thereafter. We use the following strategy to derive predictions for the three methods: MM: fitting the mechanistic model to the initial treatment data only and further simulating the future time course, GLM: feeding the explicit features of the initial time-course (see Materials and Methods) into a GLM classifier and NN: using an end-to-end learning approach with a neural network model applied to the initial time-course, which has been trained previously on an annotated reference data set (Fig. 1c).

Next, we analyzed how well the different approaches (MM, GLM, NN) can predict the outcome for the artificial patient data and how model performance changes with varying data quality (Fig. 1b,c, and Experimental Procedures). The results of the 10-fold cross-validation of the model performance are depicted in Fig. 2. As expected, the prediction accuracy (see Materials and Methods) declines for all approaches when the data quality decreases. We point out that the decrease in data quality differs between use-cases and models. In the case of AML, the introduction of sparsity leads to a relatively sharp drop in model performance. This drop illustrates the strong dependency on the number of measurements per time series: as in the given patient we only have a median of 4 measurements in the SN and AP data, compared to 39 weekly measurements in the dense data set (D) set. In line with this argument, we observe a more gradual decline in performance when comparing the effect of introducing noise and sparsity in the CML case. Here, we face a median of 25 measurements in the SN and AP data, compared to 93 monthly measurements in the dense data (D).

**Fig. 2:**
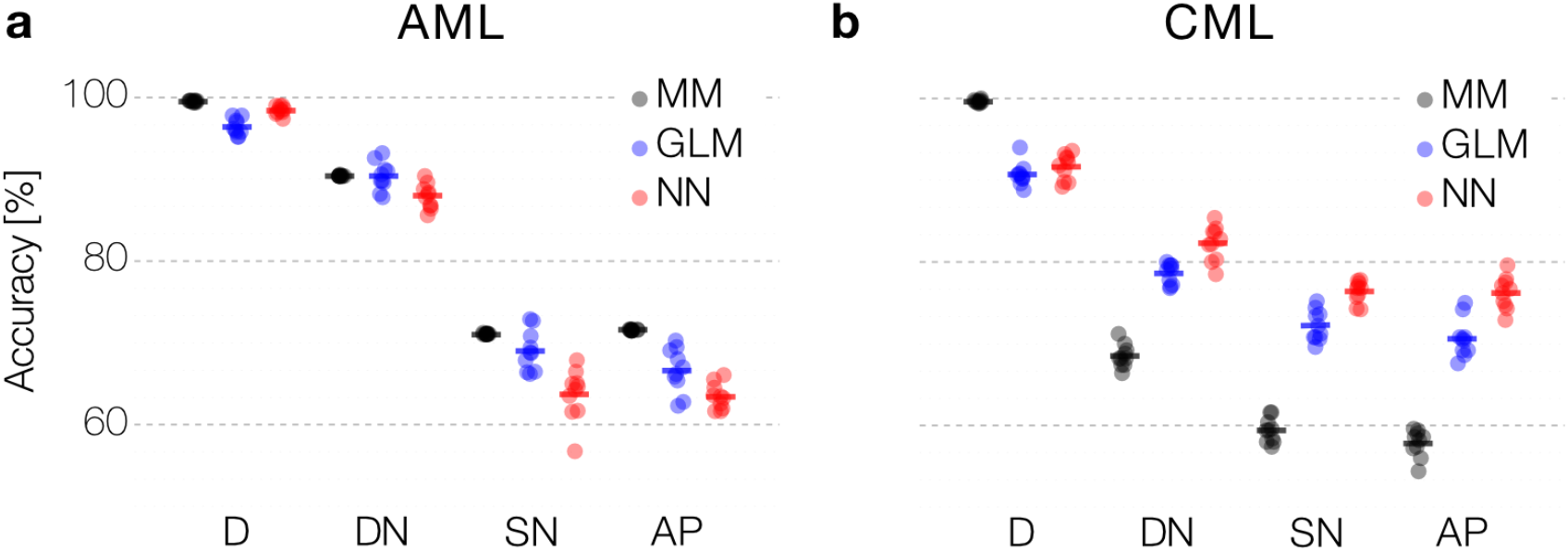
Prediction accuracy across data quality and computational models: (a, b) Comparison of performance between mechanistic model (MM), generalized linear model (GLM) and neural network (NN) to predict relapse in synthetic data for AML (a) and CML (b) using 10-fold cross-validation. Data quality gradually decreases from fully sampled, noise-free data (D), to noisy (DN), sparse and noisy (SN), and artificial patient data (AP) (see main text for details).

Interestingly, the difference in model performance is not consistent across the two use-cases. For the sparser AML data, all models perform similarly on the dense (D) and noisy data (DN). However, when introducing more sparsity into the data, a mechanistic model performs more robustly than the generic NN model (a difference in the accuracy of 6.3 and 7.4 percentage points for the SN and AP) and the GLM model performance is in between MM and NN. This result reflects the importance of introducing prior knowledge (or inductive bias) when dealing with very few data (Fig. 2a).

We observe a different situation in the CML case. Here, the prediction accuracy for the mechanistic model drops down substantially more compared to the statistical GLM model and the generic NN when data quality decreases (a difference in accuracy between MM and NN of 19.7% for SN and 19.8% for AP, respectively). We recall that the noise-free data (D) was generated by the very same mechanistic model (compare Fig. 2b). The high prediction accuracy for this data indicates that the correct (generative) MM can truly be identified. However, given the higher number of free parameters (n=7) in the CML case, a reduction of data quality (either resulting from noisy or sparse measurements) more strongly affects the identifiability of the correct MM, while the GLM and the NN appear more robust.

Focusing on the artificial patient samples (AP), which best mimic the available patient data sets, the suggested models reach an accuracy of up to 70% (compare Figure 2a,b). These findings shows that predictive computational methods can indeed support risk assessment in myeloid leukemias based on nontrivial patterns in time series data obtained during treatment. However, the resulting prediction accuracy might not adhere to the expected standards for clinical decision support. Our systematic analysis shows how data characteristics, in particular the measurement schedule, effects the performance. Data scarcity and limited accuracy of available measurements per patient appears as a limiting factor for the overall prediction accuracy for relapse occurrence. Given those constrains on the data side, we are skeptical that structural changes to the computational methods (e.g. by refining the neural network architecture) can substantially improve the overall performance. However, below we outline the potential in optimizing the measurement process to yield more informative sampling schemes.

### Refined measurement and treatment schemes lead to improved prediction accuracies

We demonstrated that a significant limitation for the prediction accuracy results from the sparsity of the available data, in particular for the case of AML. Here, molecular diagnostics and especially bone marrow aspirates are limited resources in the clinical setting. As only increasing the sampling frequency is not an option in many cases, we wondered whether an optimized timing of the measurements could lead to better predictions while the overall number of measurements remains the same. To investigate this question, we created an additional set of artificial patients (AS) with consistent measurement intervals during the nine-month treatment period (i.e. the first day of each therapy cycle and every six weeks during the treatment-free phase). This typically results in 4 to 8 (median = 7) measurements per patient, which is only a moderate increase to the reported median of 5 measurements in the clinical sample. Fig. 3a indicates that for this amended sampling regimen, we can already increase the accuracy of all prediction approaches (MM and NN by up to 12%, less pronounced for GLM). This finding strongly suggests that an adapted sampling scheme can considerably contribute to better relapse predictions, e.g. using methods from an optimal experimental design [33–36].

**Fig. 3:**
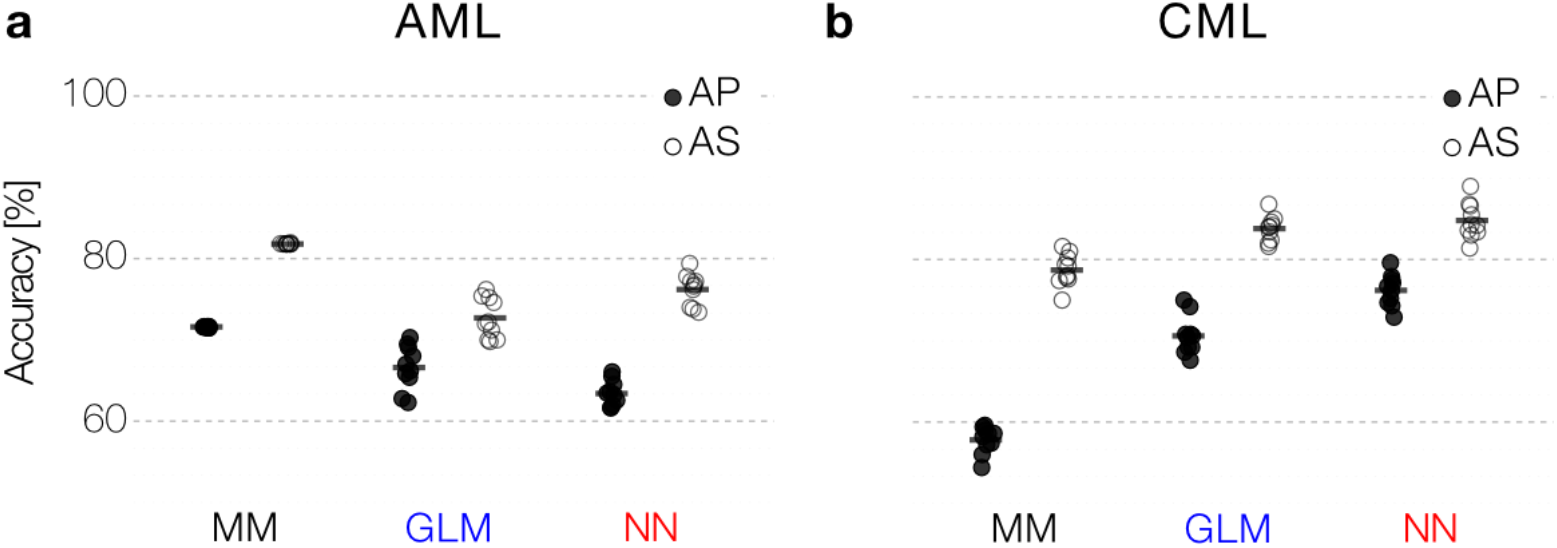
Dedicated measurement schemes: (a, b) A dedicated measurement scheme (AS) improves prediction performance with the same number of data points for all models compared to the AP data both for AML (a) and CML (b) data.

Owing to the establishment of regular BCR-ABL measurements in TKI-treated CML patients, available time courses are usually sufficient to monitor treatment response and remission status. It is still controversial, to which extend treatment free remission correlates with the observed time course of initial response [25,31]. However, results from the DESTINY trial [37] suggest that dynamics of BCR-ABL increase during TKI dose reduction correlates with the remission status after treatment cessation [21]. The DESTINY trial differs from other TKI stop trials as patients in molecular remission reduced their TKI dose to 50% of the original dose for 12 months before TKI was finally stopped [37]. Motivated by this study, we simulated a corresponding data set in which a 12-month dose reduction is explicitly added to the model simulation (AS dataset). Training the prediction approaches to explicitly integrate this additional 12 month perturbation period, we found a substantial increase in the prediction accuracy of up to 19.1% (Fig. 3b). We argue that probing the system’s response to perturbation (such as dose reduction) provides additional information about control mechanisms that cannot be obtained from ongoing monotherapy [19,21,31].

Our analysis demonstrates that optimized measurement schedules or systematic treatment alterations can substantially improve the accuracy of relapse predictions.

## Discussion

We showed that qualitatively different computational approaches, ranging from machine learning approaches to mechanistic models, are in principle suited to support relapse prediction based on time-series data of leukemia remission levels. To this end, we employed simulated time course data generated by mechanistic mathematical models, which we previously developed to describe disease and treatment dynamics in CML and AML. It is the advantage of this approach that we obtain highly controlled, although idealized, remission curves as a reference set from which we can abstract different levels of sampling density and measurement error. The simulated data allows us to refer to the *ground truth* of the underlying generative model. Using this artificial reference data, we could demonstrate that data quality in terms of measurement frequency and measurement error has a more substantial influence on the accuracy of the prediction than the employed prediction method, which is particularly evident in the AML data. Our results for the CML case indicate that fitting a more complex mechanistic model (in terms of the number of model parameters) to noisy data yields a greater uncertainty compared to a statistical predictor like a GLM or a NN.

Our analysis illustrates that generic methods, such as NN work well for the prediction of disease recurrence if frequent measurements are available (as in the CML data). For diseases with sparse measurements and limited data on the other hand (exemplified in the AML data), neural networks (and representation learning in general) is less suited for identifying the critical factors underlying the disease dynamics. In such cases, it is beneficial to incorporate prior knowledge to yield better predictions using either mechanistic models of the disease, if available, or statistical approaches based on explicit (phenomenological) features. In our current study, we used a long-short-term-memory (LSTM) NN as the standard approach for analyzing sequential data. An interesting next step is to assess if more complex neural network models [38,39] can even improve the LSTM results, although we suspect that data quality is the major limiting factor.

Regardless of the exact choice for a predictive computational method, our study indicates that the optimization of measurement schemes and clinical protocols is a promising strategy to improve the overall prediction accuracy without necessarily requiring more measurements per patients. In our predictions for AML recurrence, we could reach a level of accuracy of about 80% for the prognosis of relapse occurrence within two years after diagnosis. This result would already exceed the prediction accuracy for relapse-free survival after 12 months in the study by [18]. As our results are based on synthetic data, this comparison should be treated with caution and needs to be validated using independent clinical data obtained in a comparable context. Still, our findings indicate that standardized measurement schedules add critical leverage to improve the ability for predicting relapse no matter what computational methods are used. Our artificial measurement schemes indicated a clear improvement, while we did not even apply formal optimization criteria to obtain most suitable regimes that maximizes accuracy while minimizing the number of measurements. This finding opens a clear perspective for future research on optimized measurement strategies that balance a maximized gain of information from clinical data with an economical use of resources. We argue that such refined schedules can contribute to reaching a level of prediction accuracy, which indeed supports clinical decision making.

In this work, we focused on the accuracy of relapse prediction employing three different, prototypic computational approaches working on time-series data. However, their implementation in a decision-making context also requires an intuitive understanding of how the method works. Although NN do not require any prior knowledge and can achieve excellent prediction accuracies, it is not trivial to identify which aspects of the data are causative for a particular prediction [40,41]. In other words, the “black box” nature of NN does intrinsically not reveal the key features of the data on which a decision is based. There is a general, ongoing scientific discussion whether this intrinsic limitation of NN should prevent its application for particular questions, especially in health care [42,43]. Currently, decision-makers and regulatory authorities hardly consider such methods for integration into clinical routines, although this might change in the future. MM represent the other side of the “interpretability spectrum” as they superimpose a principal understanding of the underlying interactions onto the final observations. It appears tempting to favor this type of approach. However, it comes with other limitations: such models are highly specific and not easily transferable to other disease entities, and it cannot be guaranteed that all essential interactions are indeed mapped (compare [23]). GLMs represent a middle ground and balance several aspects of NN and MM approaches. They can be helpful if detailed mechanistic knowledge is missing while important features of the response characteristics can readily be named, estimated and also interpreted.

However, their overall performance depends strongly on the choice of those hand-crafted features and is also vulnerable to missing critical aspects.

The increasing availability of diagnostic methods to track molecular remission in different cancer types over extended time periods will establish a rich data source to explore further how this dynamic information can be correlated with the future course of treatment and disease [19]. Obtaining a systematic understanding of how different computational methods can be used to exploit this data is of crucial importance to provide usable predictions and potentially integrate them into decision making in the clinical context.

## Data Availability

The patient data for the AML patients are published in [Hoffmann2020] and can be found here: https://doi.org/10.6084/m9.figshare.12871777.v1
The CML patient data will be provided upon request to the correspondent author (Ingmar Glauche).
Source code is available at https://zenodo.org/record/4293490#.X8DznMtKg-Q

https://doi.org/10.6084/m9.figshare.12871777.v1

## Data and Code Availability

Patient data for AML patients was published in [23] and can be found at https://doi.org/10.6084/m9.figshare.12871777.v1 The CML patient data was published in [31]. Source code is available at https://zenodo.org/record/4293490#.X8DznMtKg-Q

## Acknowledgements

This work was supported by the Technische Universität Dresden, Faculty of Medicine Carl Gustav Carus, MeDDrive grant no. 60470 to NS. We thank clinical partners for providing the patient data within the originals works [22,23,31] on which this simulation study is based. Open Access Funding by the Publication Fund of the TU Dresden.

## Author Contributions

HH generated the AML data, carried out the analyses with the mechanistic model and the GLM for the AML use-case, conducted the numerical studies with the neural networks and drafted the manuscript. CB generated the CML data, carried out the analyses with the mechanistic model and the GLM for the CML use-case, and drafted the manuscript. TZ and AG were involved in creating a concept and a corresponding prototype. They critically revised the manuscript. SW, SN and NS revised the implementation of the neural network and improved the parametrization. EK contributed to the parameterization of the CML models. IG and IR contributed conceptually to the development and application of the GLM and the mechanistic models. IR critically revised the manuscript. IG and NS gave feedback during the analyses and drafted the manuscript.

## Supplementary Figures

**Supplementary Figure 1:**
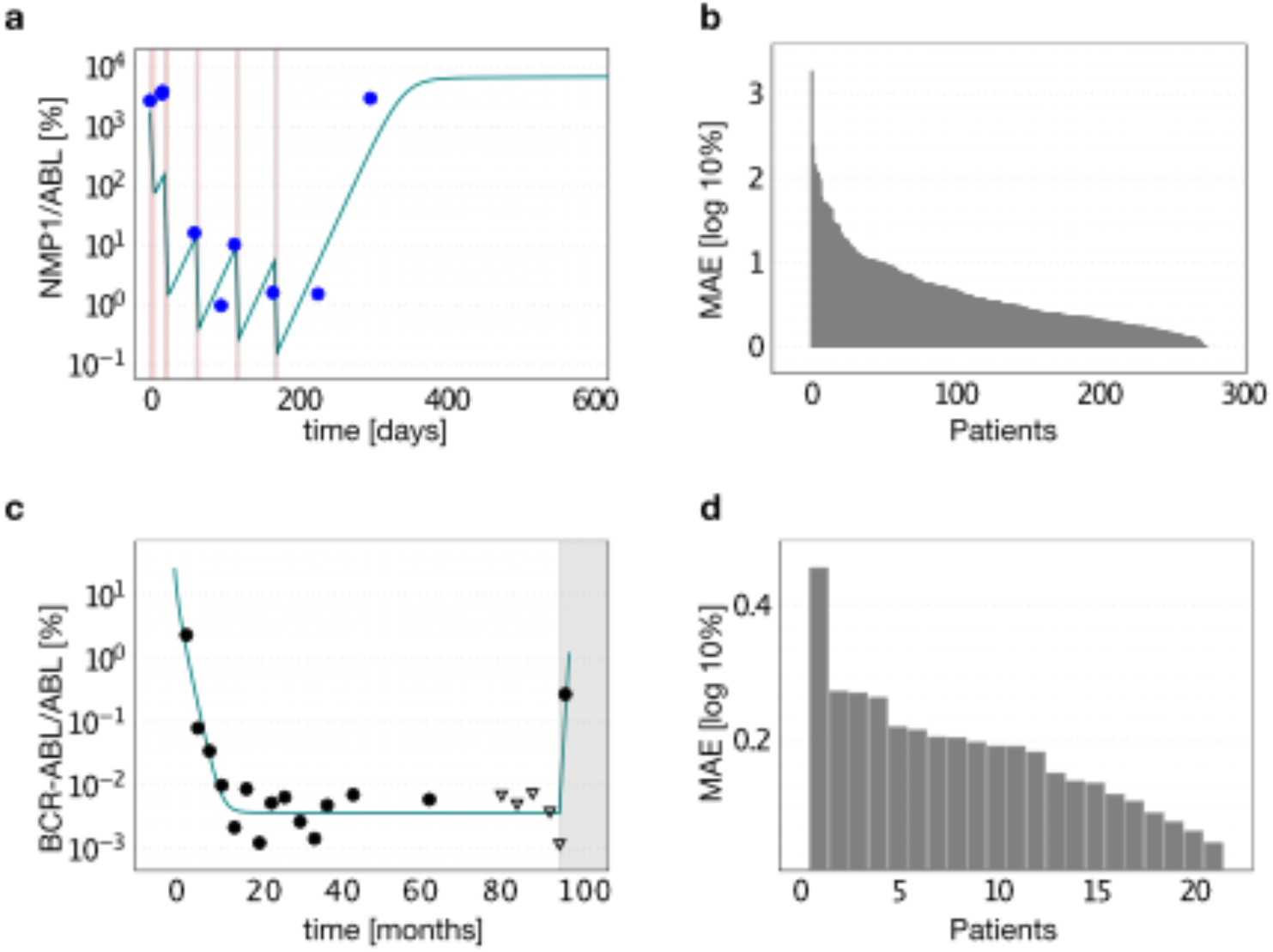
Mechanistic model fit to patient data: (a) Example time-course of an AML patient (measured in terms of NPM1-mut abundance relative to reference gene ABL; blue dots) from start of chemotherapy at time point 0 until molecular relapse and the respective model fit (solid line; leukemic burden, rescaled by a factor 100 to match the clinical NPM1-mut/ABL ratios [23]). Red lines indicate time of chemotherapy administration. (b) Mean absolute error (MAE) for the fit of the mechanistic model to all 275 AML patients time-courses. (c) Example time-course of a CML patient (measured in terms of BCR-ABL/ABL abundance; black dots; triangles indicate undetectable BCR-ABL levels with the corresponding detection threshold) from start of TKI treatment at time point 0 until disease recurrence after treatment stop (grey region) and respective model fit (solid line). (d) MAE of all 21 fitted CML patients.

**Supplementary Figure 2:**
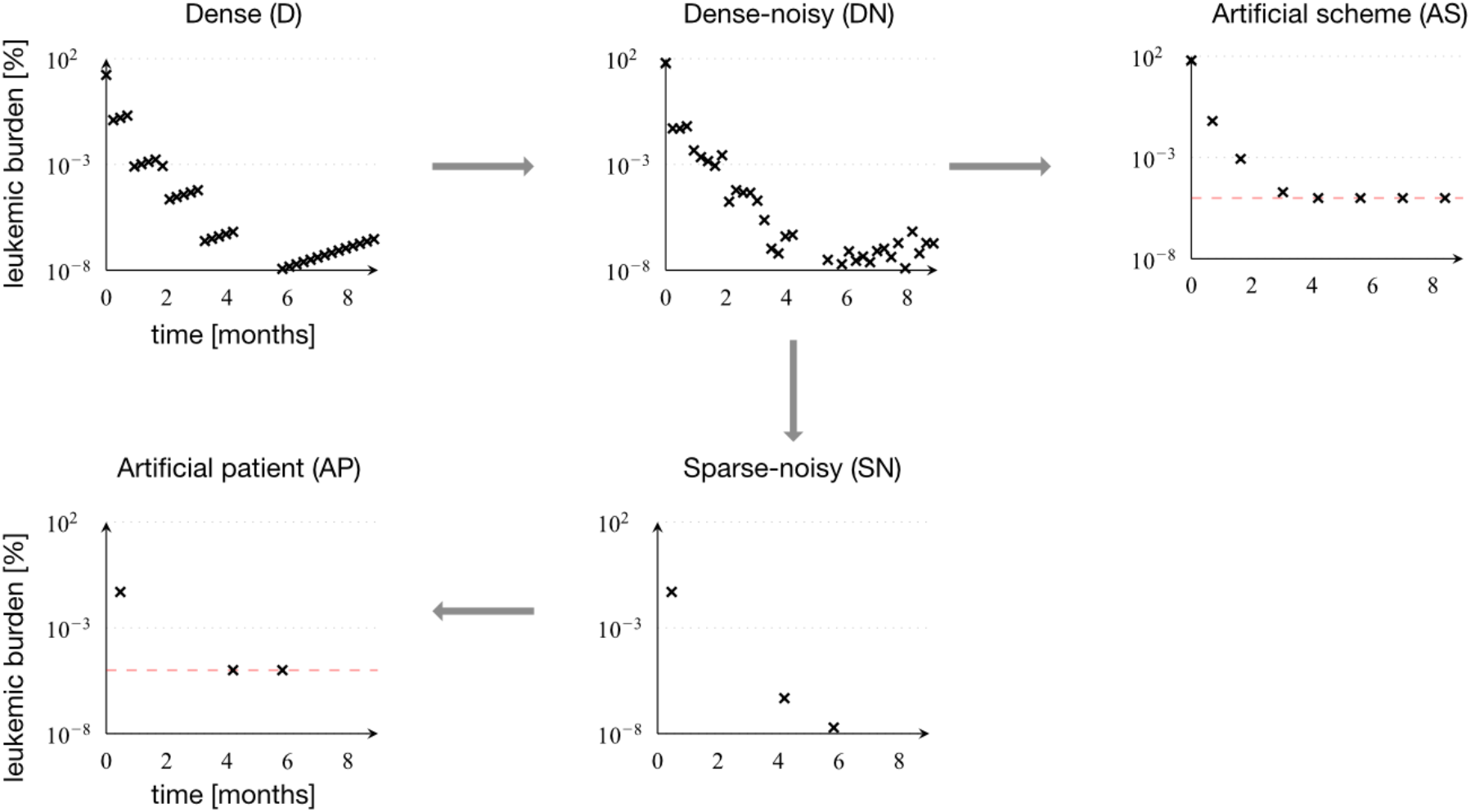
Generation of the artificial AML data sets: We use a sample patient for which we obtain weekly and precise measurements, referred to as *dense* data (D). Adding a technical, normally distributed noise to each measurement on the log-scale, we obtain *dense-noisy* data (DN). *Sparse-noisy* data (SN) was generated from the DN data set, by reducing the number of data points to meet the measurement frequency in real patients. *Artificial patient* data (AP) is the data set most similar to the real patient data, which differs from the SN data set only by the inclusion of a detection limit (dashed red line), as it is found in the real data. *Artificial scheme* data (AS) is a data set, close to real data, with a measurement scheme, where measurements are made at the end of each chemotherapy cycle and every 6 weeks afterwards.

**Supplementary Figure 3:**
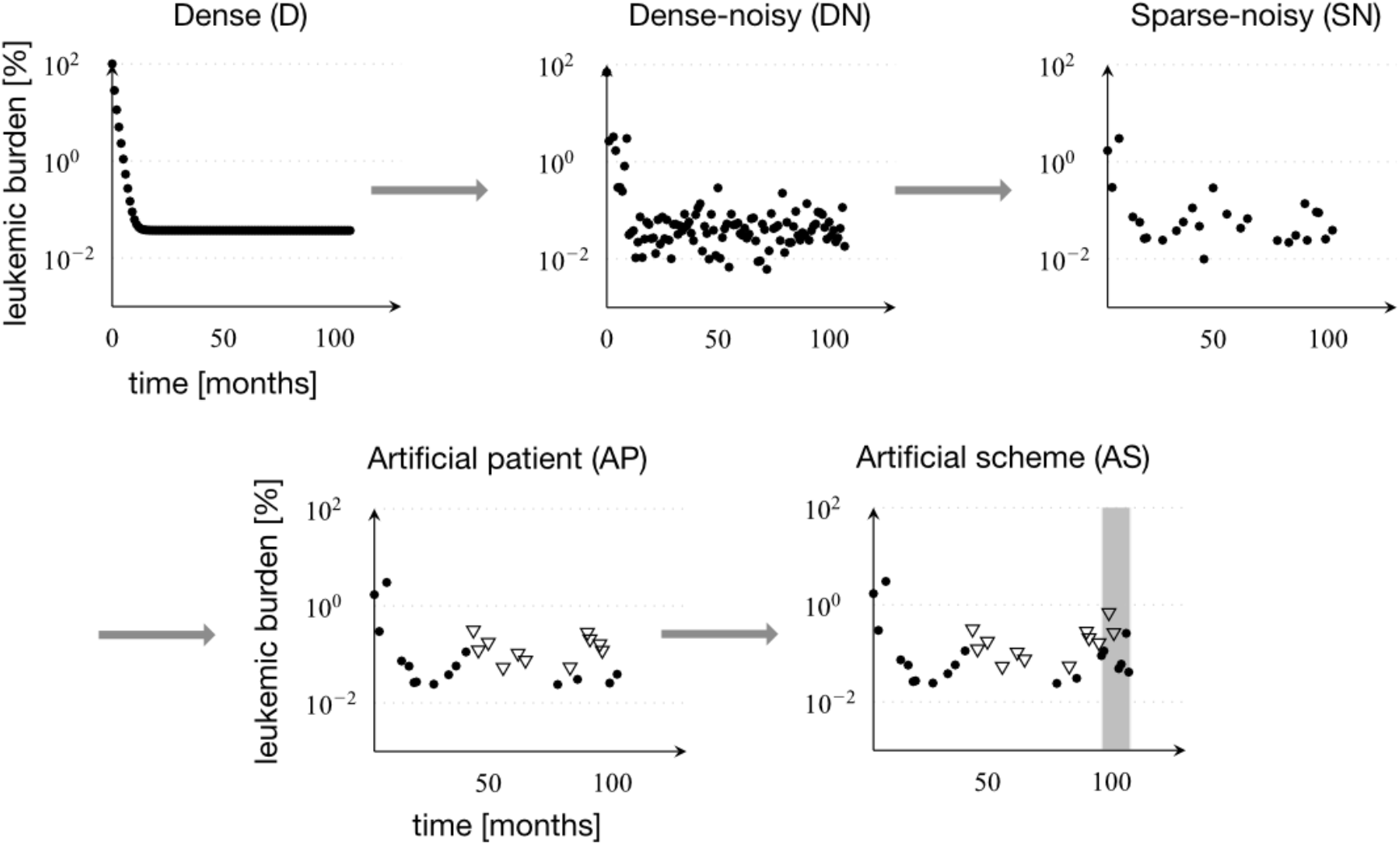
Overview of artificial CML data sets: *Dense* data (D) was simulated with monthly exact measurements. *Dense-noisy* data (DN) was obtained by adding normally distributed noise to each measurement. *Sparse-noisy* data (SN) was generated from the DN data set, by reducing the number of data points to meet the measurement frequency in real patients. *Artificial-Patient* data (AP) is the data set most similar to the real patient data, which differs from the SN data set only by the inclusion of a detection limit, as it is found in the real data. *Artificial scheme* data (AS) is a data set, close to real data, with an additional 12-month period of half-dose TKI treatment (shown in grey).

**Supplementary Figure 4:**
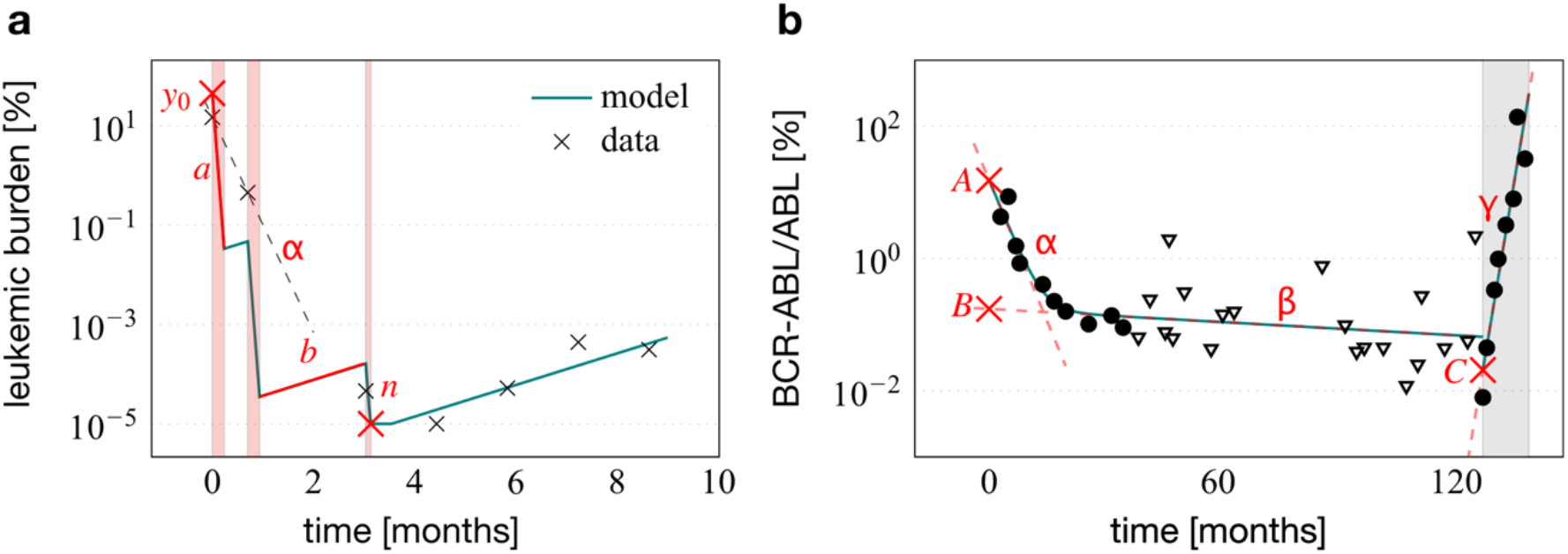
Derived features of the time-courses: (a) Features describing AML time courses: *y*_*0*_ the leukemic burden at diagnosis, *a* the decreasing slope during treatment cycles, *b* the increasing slope in treatment free intervals (where *y*_*0*_, *a* and *b* are obtained from a segmented regression approach), *α* the overall decreasing slope during treatment (shown as dashed line, separately fitted to the measurements) and *n* the minimal leukemic burden after treatment. (b) Features describing CML time courses: *A, B* and *C* being the intercepts of the straight lines fitted to the first and the second part of the bi-exponential approximation and to the increase of the leukemic burden during half-dose periods, respectively. *α, β* and *γ* are the respective slopes.

**Supplementary Figure 5:**
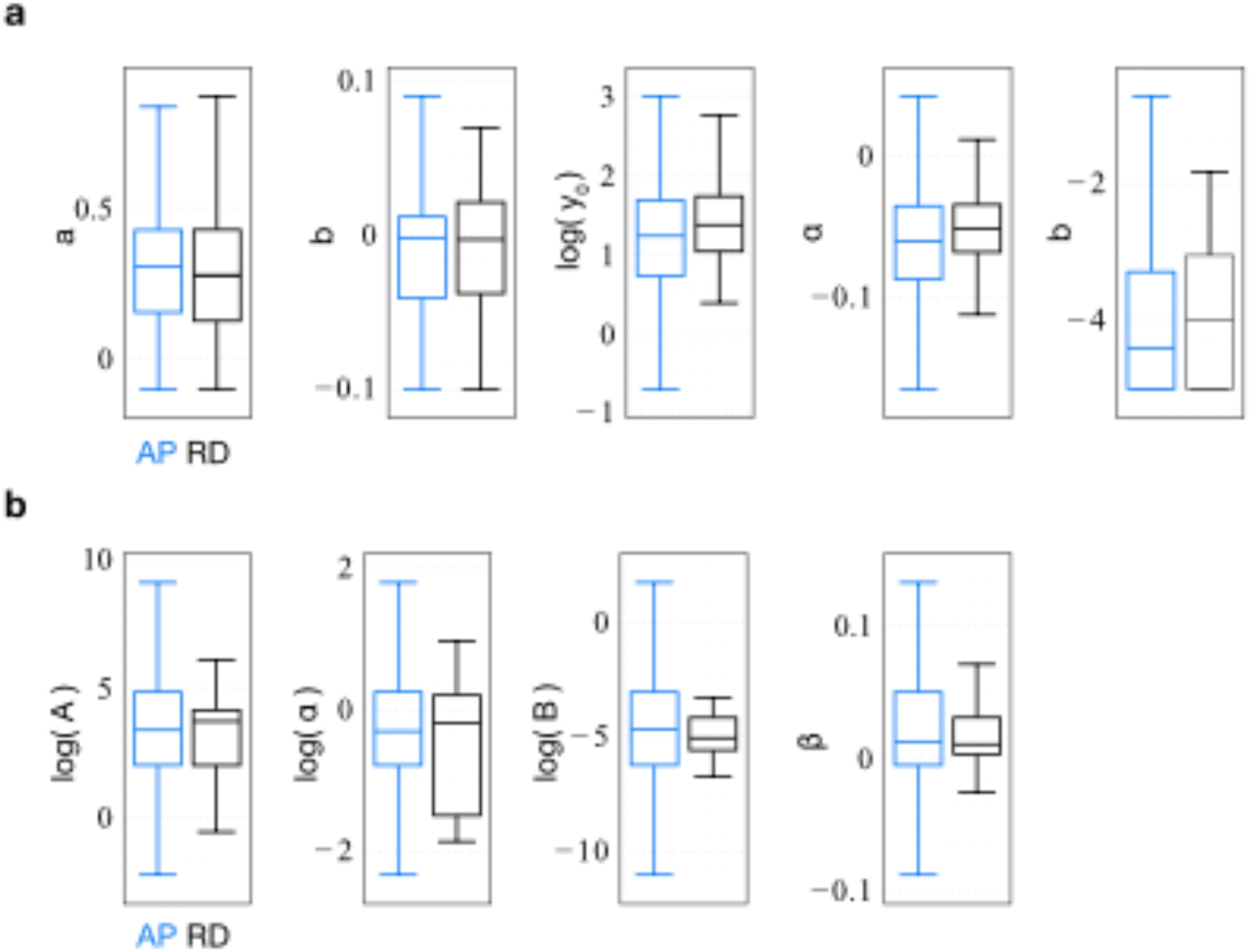
Similarity of artificial patients and real patients: a distribution comparison of statistical parameters: Comparison of distribution of parameters describing the course characteristics between artificial patient data (AP, blue) and real data (RD, black). (a) Parameters characterizing AML response: a - decreasing slope during chemotherapy cycle, *b* - increasing slope during treatment-free periods, *y*_*0*_ -initial burden on log scale, *α* - elimination slope, *n* - minimal measured leukemia burden after primary treatment. (b) Parameters characterizing CML response: the intercepts *A* and *B* (on a log scale) as well as the slope parameters *α* (on log scale) and *β*.

**Supplementary Figure 6:**
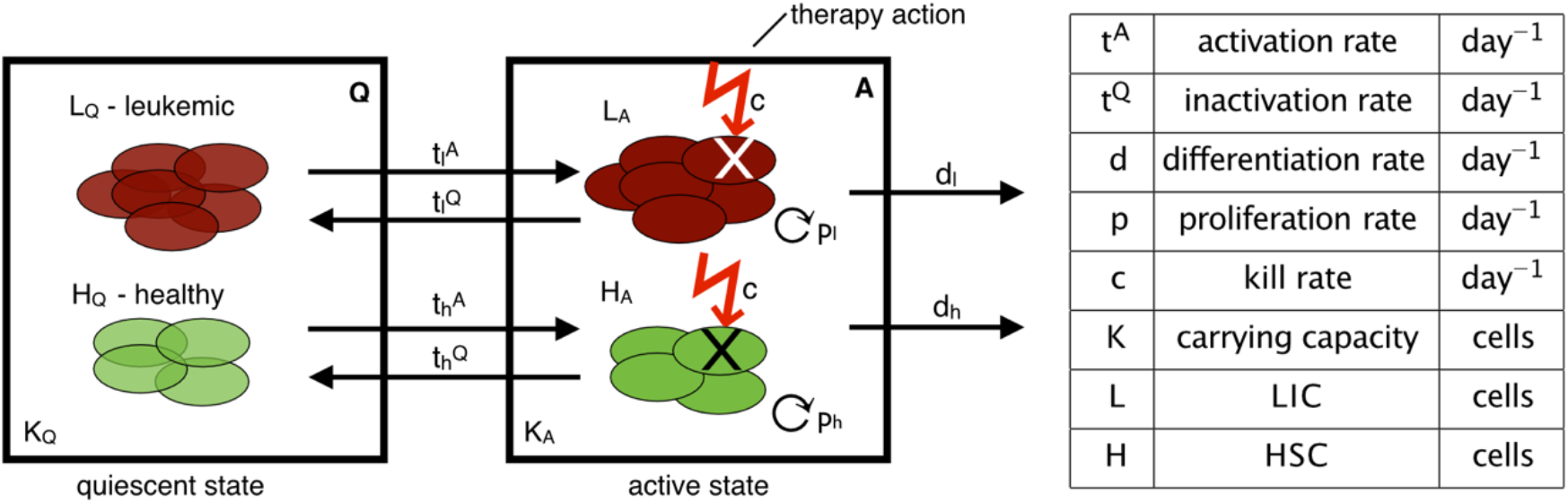
Schematic overview of the mathematical model for AML: Both, leukemic *L* and healthy *H* stem cells can reversibly change between two states (according to the rates *t)*: the quiescent state Q with carrying capacity *K*_*Q*_ and the active state A with carrying capacity *K*_*A*_. Cells in A undergo proliferation with rate *p*, differentiation with rate *d* and are subject to chemotherapy with kill rate *c*.

**Supplementary Figure 7:**
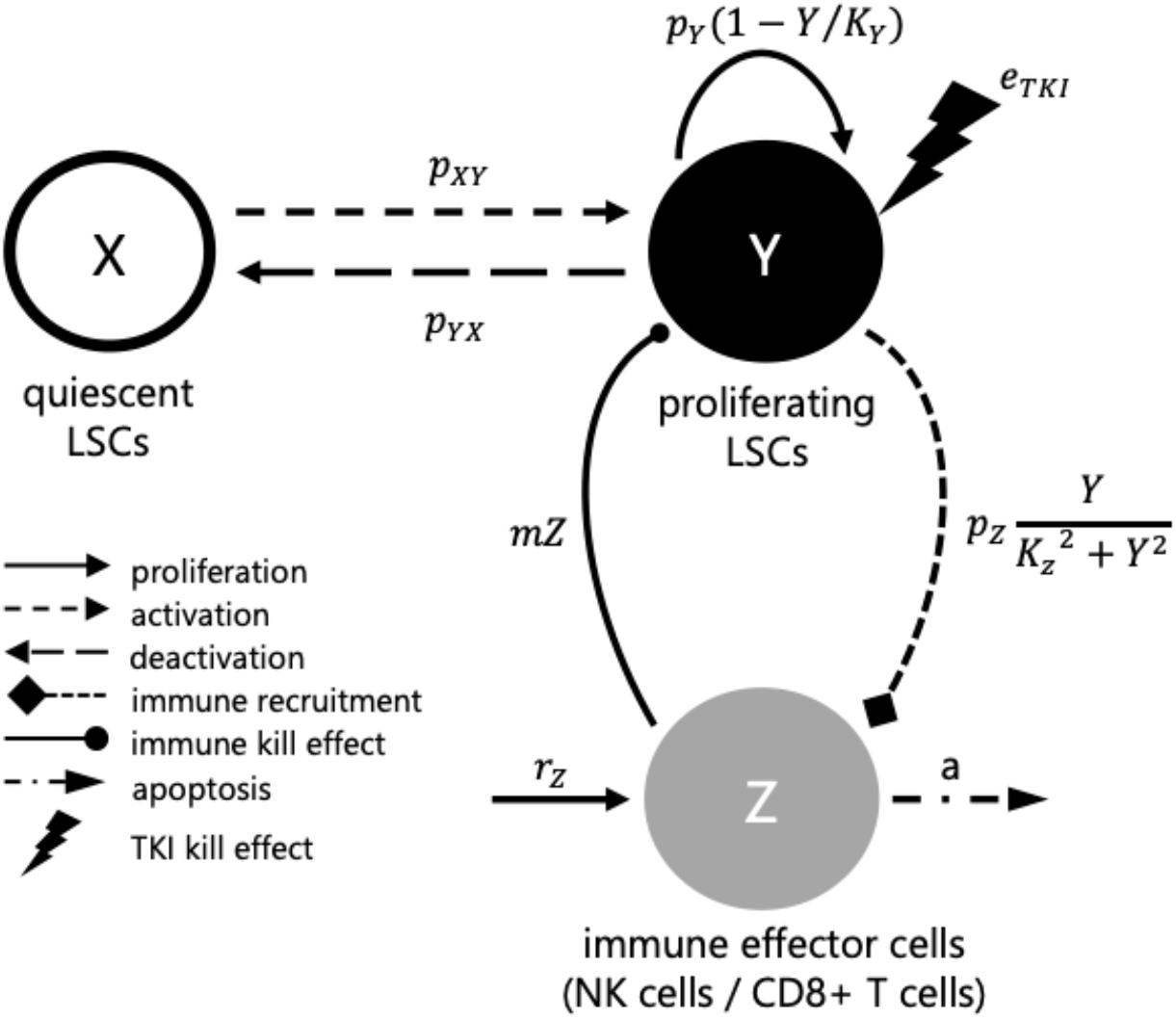
Schematic overview of the mathematical model for CML:. Leukemic stem cells (LSC) can reversibly change between two states X and Y (according to the rates *p*_*XY*_ and *p*_*YX*_, respectively): X defines the quiescent, non-replicating cells, Y defines the active, proliferating cells. LSC in Y proliferate according to a logistic growth model with maximal proliferation rate *p*_*Y*_ and carrying capacity *K*_*Y*_. The TKI-effect is described by a constant rate *e*_*TKI*_ affecting the leukemic cells in Y. Immune cells in Z are activated by cells in Y (immune recruitment), following an immune window approach (see Supplementary Text). At the same time the immune cells kill proportional target cell in Y. Immune cells in Z are generated with rate *r*_*z*_ and decay with rate *a*.

## Supplementary Text

### AML

#### Clinical data

To be able to generate data with the mechanistic AML models that were as similar as possible to real clinical data, we used a patient data set for comparison. The data published together with the mechanistic model was used [23]. 61 of these 275 patients relapsed within two years after therapy initiation. The data consists of time course measurements of the relative tumor load of NPM1-mutated patients, by qPCR measurements of the amount of NPM1-mut transcripts relative to the amount of the reference genes transcripts (ABL).

#### Mechanistic model

Our mechanistic model of the molecular disease dynamics of AML, already published in [23], describes the dynamics of healthy stem cells (*H*) and leukemia-initiating cells (*L*) in the bone marrow of an AML patient (schematic overview in Figure S6). Each cell can adopt one of the two differential states: a quiescent state (Q) and an active state (A). The cells can reversibly switch between states with transition rates *t*^*A*^_*l/h*_ and *t*^*Q*^_*l/h*_. Cells in active state (*H /L*) proliferate with proliferation rate *p*_*l/h*_ and are sensitive to chemotherapeutic treatment with the kill rate *c*. Active cells can also differentiate into other states and therefore leave the two observed states. Individual chemotherapy schedules as well as patient-specific transition rates from quiescent to active state of the leukemic cells (*t*_*l*_^*A*^) and proliferative potential of leukemic cells (*p*_*L*_) are taken into account to adapt the model to individual patients. The model equation are as follows:

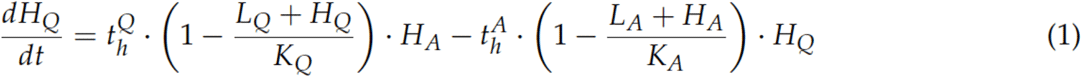

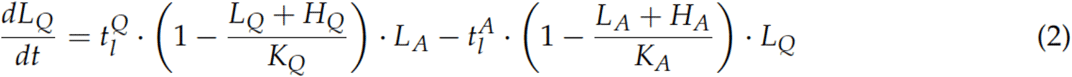

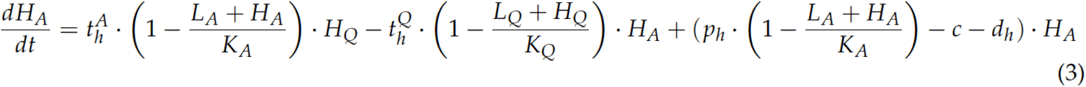

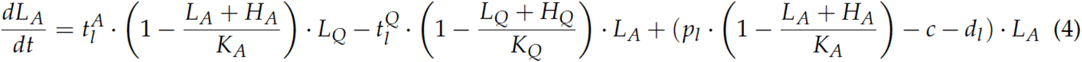

For individual patient fitting we minimized the residual error between model and data using the *MultiStart* function and the *fmincon SQP* algorithm in MATLAB. The search space for parameter fitting was set as follows: for *t*_*l*_^*A*^ between 0.01 and 0.99 and for *p*_*l*_between 0.04 and 0.2. The model was initialized using the steady state of the model with not leukemia advantage (meaning all parameters are the same for leukemic and healthy cells) and starting with 60% healthy and 40% leukemic cells in each compartment.

The other parameter values were fixed as follows: *K*_*A*_ = *K*_*Q*_ = 105, *t*_*h*_^*A*^ = 0.01, *t*_*h*_^*Q*^ = 0.2, *t*_*l*_^*Q*^ = 0.002, *p*_*h*_ = *0*.*04, d*_*h*_ = *d*_*l*_*= 0*.*033, c = 0*.*99*.

#### Artificial data

For data generation, we took the chemotherapy information obtained from real patients (i.e. timing and number of cycles) and combined them with fitted parameters from the AML patient set [23]. The parameters were slightly varied by adding a white noise (standard normal distributed with *μ*_*P*_ *= 0* and *σ*_*P*_ *= 0*.*01*). The following 5 data sets with 5000 time-courses each, half of them relapsing, half of them non-relapsing within two years, and a measurement period of 9 months were generated (Supplementary Table 1):

- Dense (D) data: one measure per week starting with the first day of chemotherapy.
- Dense-noisy (DN) data: added white noise (*μ*_*D*_ *= 0, σ*_*D*_ *= 0*.*5*) to the log-scaled data from D.
- Sparse-noisy (SN) data: Measurement frequency was adjusted to the number and intervals of measurements in the real data. Therefore, the real data was divided into three intervals (first 3 months, second 3 months and third 3 months) and for each interval the number of measurements was counted and randomly sampled for each artificial patient. From the DN data, a weighted sample of this number of measurements was then taken using the histogram of the time points in each interval to weight the sampling.
- Artificial patient (AP) data: all values of SN below the detection limit of -5 [log10] were set to -5 [log10].
- Artificial scheme (AS) data: using DN measurement time points at the beginning of each therapy cycle and every 6 weeks thereafter. All values below detection limit were set to -5 [log10].

The leukemic burden, which is the main readout of the model, is calculated as the relative fraction of leukemic cells in both states with respect to the overall number of all cells in these states, i.e.:

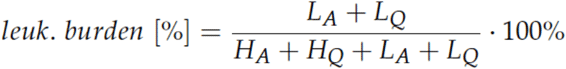

The leukemic burden is rescaled by a factor of 100 to compare it to the patient’s NPM1/ABL measurements [23]. Only patients which reached remission after therapy and did not relapse within the first 9 months were included in the data set. If the leukemic burden exceeded the relapse threshold of 1% at two years after treatment start, the time series was classified as a relapse.

**Supplementary Table 1:** Number of patients and average number of measurements for all AML datasets. The reduced number of patients in SN, AP results from exclusion of cases where no measurements were scheduled from our algorithm.

|  | number of patients | average measurements per patient |
| --- | --- | --- |
| **clinical data** | 275 | 3.8 |
| **D** | 5000 | 39 |
| **DN** | 5000 | 39 |
| **SN** | 4973 | 3.8 |
| **AP** | 4973 | 3.8 |
| **AS** | 5000 | 7 |

#### Generalized linear model

For training a generalized linear model (GLM), in more depth a logistic regression classifier, we derived 5 features. Two were taken from the previous description of the main characteristics of the time course in AML [22]: the elimination slope α, giving a measure of the decrease of leukemic burden during therapy, until the relapse threshold of 10^−3^ is reached and the minimal leukemic burden after therapy (n). The other three features were derived from a simple model describing the time course with a starting point (y_0_) at the beginning of treatment, a linear decrease with slope *a* of the leukemic burden during the time of treatment and a linear increase with slope *b* during treatment-free periods (see Suppl. Figure 4A). The starting points (y_0_) and the two slopes (*a* and *b*), together with the two characteristics were then handed to the GLM. When fitting the GLM using the parameters it became clear that two of them could be left out without losing accuracy. Therefore, the final GLM was fitted to predict the relapse based only on the minimal leukemic burden after therapy (*n*), the decreasing slope during therapy (*a*) and the increasing slope during treatment free periods (*b*). A 10-fold cross validation was used to estimate the variation of the estimated accuracy.

#### Neural Network

The neural network consists of a bidirectional long-short-term-memory (LSTM) layer with 32 hidden units (using ReLU activation), followed by a fully connected layer with 64 nodes with a ReLU activation and a single, sigmoid output layer.

We use a vector of log10 values of NPM1/ABL values as input. Missing values (i.e. no measurement available at this particular time point) were encoded as -1 and a respective masking-layer was introduced to the network. For the non-detectable datapoints, we used the detection limit for the clinical data (see above). The time points of chemotherapy were given as a second channel input in the case of AML.

To prevent overfitting, 10-fold cross validation was used. We saved the model with the highest validation (not training) accuracy. The entire training process was repeated 10 times on each dataset, to report a more robust estimate of the network performance as we regularly observed numerical instabilities in the training process leading to failure of model fitting. Networks were trained with binary cross-entropy loss using the Adam optimizer with a learning rate of 10^−3^ and a learning rate decay of 10^−5^. The size of the validation set was chosen as 15 % of the training set. We used a batch size of 128 and trained for 100 epochs using an early stopping checkpoint to stop the training if the validation loss did not decrease in 20 epochs.

### CML

#### Clinical data

To generate in-silico CML patients as similar to real patients as possible, we utilized a cohort of 21 CML patients based on [31].

Each measurement can be either a detectable or an undetectable value. If a measure is detectable a corresponding BCR-ABL1 ratio on a log scale (LRATIO) is given. An undetectable measure is defined by a quantification limit (lQL) dependent on how much of the reference gene ABL1 was found in the sample. The more ABL1 is in the sample, the lower is the quantification limit.

For CML, typically a biphasic course in the time series is observed. At first, rapid decline symbolizes the fast clearing of the majority of tumor cells. This phase usually takes between 6 and 12 months. After this initial phase, a second phase with a moderate decline begins, symbolizing the slow tumor degradation in bone marrow. During the treatment the tumor load measurable in blood becomes lower, thus the number of non-detectable measures increases over time.

#### Mechanistic model

The mechanistic model simulating the CML cells consists of 3 compartments (Supplementary Figure 7). X defines the quiescent, non-replicating cells, Y defines the active, proliferating cells and Z defines the immune cells specific to the CML cells described by X and Y. Cells change from a quiescent state into an active state and vice versa with certain probabilities defined by the rates *p*_*XY*_ and *p*_*YX*_, respectively. Furthermore, Y cells proliferate with a logistic growth defined by a maximal rate *p*_*Y*_. The TKI-effect is described by a constant rate TKI killing the corresponding proportion of leukemic cells in Y. Immune cells in Z get activated by the number of active cells in Y. At the same time the immune cells kill proportional cells from Y depending on its number. However, the activation function of Z, depending on Y, defines a so-called immune window. In other words, depending on the parametrization immune effector cells are suppressed on very high and very low numbers of active leukemic cells. In between the immune cells rise and get effective [31]. The model is implemented in *Julia*. For this model we are using the default Julia ODE solver configured like follows: We assume a non-stiff problem and apply the *Tsit5* algorithm. During solving, an auto-detection for stiffness is applied and in case stiffness is detected, the solver is switched to the *Rosenbrock23* solver.

#### Mechanistic parameters

To fit the model to clinical and in-silico data, we used the following model parameters:

- maximum activation rate of Z (*p*_*Z*_)
- immune window suppressing constant (*K*_*Z*_)
- cell switch rate from X to Y (*p*_*XY*_)
- cell switch rate from Y to X (*p*_*YX*_)
- maximum activation rate of Y (*p*_*Y*_)
- TKI kill rate (*e*_*TKI*_)
- initial tumor burden (*initRatio*)

The following model parameters were fixed among all mechanistic simulations:

- maximum number of tumor cells in Y (*K*_*Y*_ = 10^6^)
- kill rate of active leukemic stem cells by immune cells (*m* = 10^−4^)
- constant additive influx of immune cells (*r*_*Z*_ = 200)
- apoptosis rate of immune cells (*a* = 2.0)

#### Fitting Mechanistic Model to Data

We fitted the mechanistic parameters of all clinical as well as in-silico patients in R using a genetic algorithm from a modified R-package *rgenoud* (version 5.8-3.0). The specific configuration and the corresponding package can be found in the repository. The fitness function is defined as the sum of the distance of all measurements with the following rules:

1. detectable measures: quadratic distance between patient data and in-silico data
2. undetectable measures: left censoring of values meaning if the simulation value is higher than the given lQL value (see Clinical Data) we use a quadratic distance, in case it is equal or lower, we use a 0-distance.

#### Artificial data

We generated the in-silico parameters using copulas to sample from the fitted patient parameters, such that we receive a highly similar correlation structure:

1. Using Copulae (R-package *copula* 1.0-0), describing a functional dependency between the marginals of multiple independent variables and their corresponding joint probability distribution, we:
  a. create a normal Copula with the correlation matrix reflecting the correlations of the empirical random variables,
  b. generate the random observations with the Copula targeting marginal uniform distributions,
  c. and transform it into the observed empirical distribution.

From this, we sampled 5000 in-silico patient parameters, whereas half are relapsing and half are non-relapsing patients (Supplementary Table 2). Additionally, we are taking the information of the properties of cessation time, measurement noise, density and frequency of undetectable values from the patient data. As properties differ at specific treatment phases, e.g. sample frequency is usually higher in the beginning, we are using the following sampling time intervals: 0-6, 6-12, 12-24, 24-36, 36-(cessation time).

1. Dense Data (D): This dataset represents the raw simulation data taking one measurement per month. All measurements are noise-less and detectable.
2. Dense-noisy Data (DN): We introduce noise, assuming it is solely represented by the distance between clinical data and the corresponding simulation fits. Therefore, we create an empirical noise distribution (END) representing the distances between clinical data and corresponding simulation fits for each interval. Next, using kernel density estimates, we add an error value for each data point in D and its correspondent interval sampling from END.
3. Sparse-noisy (SN): Using the clinical data, we calculate for each time interval a distribution of the time of the first observation (DFO) and a distribution of time intervals (DTI) by taking the differences of consecutive time points. Then, given an in-silico patient of DN and an interval, a starting time (ST) from DFO within the corresponding interval is sampled. After, the cumulative sum of sufficiently many samples from DTI is added to ST until the interval border is exceeded. Finally, only the data points of the in-silico patient from DN are kept if they are within the sampled time quantities. Doing this for all in-silico patients of DN and intervals, SN is generated from DN. All samples are taken continuously by using kernel density estimates.
4. Artificial Patient (AP): For each interval we calculate a probability that a measurement is not-detectable. These probabilities are then applied to the in-silico patients from SN to receive the final in-silico patient data.
5. Artificial Scheme (AS): We are simulating patients with a scheme inspired by the DESTINY trial (Clark et al., 2019). Therefore, we set the TKI dose in the model to 50% of the original dose 12 months before TKI cessation. To generate this AS dataset, we started all over from (1) -(4) with the only difference in the time intervals. As patients in DESTINY during dose reduction are monitored very closely, the last time interval is further divided into two parts (period from month 36 of treatment until TKI dose reduction, period of reduced TKI dose).

In all our CML simulations, we classify whether an in-silico patient relapses by simulating 10 years ahead and check whether the tumor burden is above MR1.

**Supplementary Table 2:** Overview of in-silico CML data sets in comparison with clinical data

|  | number of patients | average measurements per patient | average treatment time until cessation (months) |
| --- | --- | --- | --- |
| **clinical data** | 21 | 23.05 | 90.98 |
| **D** | 5000 | 92.51 | 92.51 |
| **DN** | 5000 | 92.51 | 92.51 |
| **SN** | 5000 | 23.19 | 92.51 |
| **AP** | 5000 | 23.19 | 92.51 |
| **AS** | 5000 | 28.80 | 92.64 |

#### Generalized linear model

To predict the relapse outcome, we trained a logistic regression classifier (GLM) using

- the first slope *α*,
- the corresponding intercept *A*,
- the second slope *β*,
- the corresponding intercept *B*,
- the standard deviation during the biexponential phase *σ*,
- the cessation time and
- the BCR-ABL1 ratio at cessation time,

whereas *α, A, β, B* and *σ* result from the corresponding biexponential fits. We used R’s default stats package to train the GLM.

In the case of the artificial scheme AS, the additional predictors were added:

- the third slope Υ during half dose,
- the corresponding intercept *C*,
- the first point in time of the half-dose and
- the BCR-ABL1 ratio at half dose time.

We allowed first-order interactions. However, resulting models were simplified based on the AIC until a minimum is reached. We used a 10-fold cross-validation.

#### Neural Network

We used mostly the same configuration as described above for the AML data. As input, we used the vector of log10 values of BCR-ABL values. In case the value was not detectable, we used the lQL (see above, Supplementary Text: CML - Clinical Data). To account for the more complex dynamics in the CML case and the higher number of data points within one time-series we increased the maximum number of epochs to 2000 with an early stopping if the validation loss does not decrease for 100 epochs.

## References

1. Rohrbacher M, Hasford J. Epidemiology of chronic myeloid leukaemia (CML). Best Pract Res Clin Haematol. Elsevier BV; 2009;22:295–302.

2. Khwaja A, Bjorkholm M, Gale RE, Levine RL, Jordan CT, Ehninger G, et al. Acute myeloid leukaemia. Nat Rev Dis Primers. 2016;2:16010.

3. Thijsen S, Schuurhuis G, van Oostveen J, Ossenkoppele G. Chronic myeloid leukemia from basics to bedside. Leukemia. Springer Nature; 1999;13:1646–74.

4. Melo JV, Barnes DJ. Chronic myeloid leukaemia as a model of disease evolution in human cancer. Nat Rev Cancer. Springer Science and Business Media LLC; 2007;7:441– 53.

5. Chereda B, Melo JV. Natural course and biology of CML. Ann Hematol. Springer Science and Business Media LLC; 2015;94 Suppl 2:S107–21.

6. Zhou H, Xu R. Leukemia stem cells: the root of chronic myeloid leukemia. Protein Cell. 2015;6:403–12.

7. Hochhaus A, Larson RA, Guilhot F, Radich JP, Branford S, Hughes TP, et al. Long-Term Outcomes of Imatinib Treatment for Chronic Myeloid Leukemia. N Engl J Med. 2017;376:917–27.

8. Bower H, Björkholm M, Dickman PW, Höglund M, Lambert PC, Andersson TM-L. Life expectancy of patients with chronic myeloid leukemia approaches the life expectancy of the general population. J Clin Oncol. 2016;34:2851–7.

9. Cerveira N, Loureiro B, Bizarro S, Correia C, Torres L, Lisboa S, et al. Discontinuation of tyrosine kinase inhibitors in CML patients in real-world clinical practice at a single institution. BMC Cancer. 2018;18:1245.

10. Mahon F-X, Réa D, Guilhot J, Guilhot F, Huguet F, Nicolini F, et al. Discontinuation of imatinib in patients with chronic myeloid leukaemia who have maintained complete molecular remission for at least 2 years: the prospective, multicentre Stop Imatinib (STIM) trial. Lancet Oncol. 2010;11:1029–35.

11. Nagafuji K, Matsumura I, Shimose T, Kawaguchi T, Kuroda J, Nakamae H, et al. Cessation of nilotinib in patients with chronic myelogenous leukemia who have maintained deep molecular responses for 2 years: a multicenter phase 2 trial, stop nilotinib (NILSt). Int J Hematol. 2019;110:675–82.

12. Cancer Genome Atlas Research Network, Ley TJ, Miller C, Ding L, Raphael BJ, Mungall AJ, et al. Genomic and epigenomic landscapes of adult de novo acute myeloid leukemia. N Engl J Med. 2013;368:2059–74.

13. Schuurhuis GJ, Heuser M, Freeman S, Béné M-C, Buccisano F, Cloos J, et al. Minimal/measurable residual disease in AML: a consensus document from the European LeukemiaNet MRD Working Party. Blood. 2018;131:1275–91.

14. Shayegi N, Kramer M, Bornhäuser M, Schaich M, Schetelig J, Platzbecker U, et al. The level of residual disease based on mutant NPM1 is an independent prognostic factor for relapse and survival in AML. Blood. American Society of Hematology; 2013;122:83–92.

15. Oliva EN, Franek J, Patel D, Zaidi O, Nehme SA, Almeida AM. The Real-World Incidence of Relapse in Acute Myeloid Leukemia (AML): A Systematic Literature Review (SLR). Blood. American Society of Hematology; 2018;132:5188–5188.

16. Döhner H, Weisdorf DJ, Bloomfield CD. Acute myeloid leukemia. N Engl J Med. New England Journal of Medicine (NEJM/MMS); 2015;373:1136–52.

17. Döhner H, Estey E, Grimwade D, Amadori S, Appelbaum FR, Büchner T, et al. Diagnosis and management of AML in adults: 2017 ELN recommendations from an international expert panel. Blood. 2017;129:424–47.

18. Othus M, Wood BL, Stirewalt DL, Estey EH, Petersdorf SH, Appelbaum FR, et al. Effect of measurable (‘minimal’) residual disease (MRD) information on prediction of relapse and survival in adult acute myeloid leukemia. Leukemia. 2016;30:2080–3.

19. Roeder I, Glauche I. Overlooking the obvious? On the potential of treatment alterations to predict patient-specific therapy response. Exp Hematol [Internet]. Elsevier; 2020; Available from: https://doi.org/10.1016/j.exphem.2020.11.006

20. Branford S, Yeung DT, Parker WT, Roberts ND, Purins L, Braley JA, et al. Prognosis for patients with CML and >10% BCR-ABL1 after 3 months of imatinib depends on the rate of BCR-ABL1 decline. Blood. American Society of Hematology; 2014;124:511–8.

21. Gottschalk A, Glauche I, Cicconi S, Clarke RE, Roeder I. Molecular dynamics during reduction of TKI dose reliably identify molecular recurrence after treatment cessation in CML. Blood. 2020;

22. Hoffmann H, Thiede C, Glauche I, Kramer M, Röllig C, Ehninger G, et al. The prognostic potential of monitoring disease dynamics in NPM1-positive acute myeloid leukemia. Leukemia. 2019;33:1531–4.

23. Hoffmann H, Thiede C, Glauche I, Bornhaeuser M, Roeder I. Differential response to cytotoxic therapy explains treatment dynamics of acute myeloid leukaemia patients: insights from a mathematical modelling approach. J R Soc Interface. 2020;17:20200091.

24. Saussele S, Richter J, Guilhot J, Gruber FX, Hjorth-Hansen H, Almeida A, et al. Discontinuation of tyrosine kinase inhibitor therapy in chronic myeloid leukaemia (EURO-SKI): a prespecified interim analysis of a prospective, multicentre, non-randomised, trial. Lancet Oncol. 2018;19:747–57.

25. Shanmuganathan N, Pagani IS, Ross DM, Park S, Yong AS, Braley JA, et al. Early BCR-ABL1 kinetics are predictive of subsequent achievement of treatment-free remission in chronic myeloid leukemia. Blood [Internet]. 2020; Available from: http://dx.doi.org/10.1182/blood.2020005514

26. Zhang X. Time series analysis and prediction by neural networks. Optim Methods Softw. Taylor & Francis; 1994;4:151–70.

27. Goodfellow I, Bengio Y, Courville A. Deep Learning. The MIT Press; 2016.

28. Fawaz HI, Forestier G, Weber J, Idoumghar L, Muller P-A. Deep learning for time series classification: a review [Internet]. arXiv [cs.LG]. 2018. Available from: http://arxiv.org/abs/1809.04356

29. McCullagh P, Nelder JA. Generalized Linear Models (Chapman & Hall/CRC Monographs on Statistics and Applied Probability). 2 edition. Chapman and Hall/CRC; 1989.

30. Lundberg SM, Lee S-I. A Unified Approach to Interpreting Model Predictions. In: Guyon I, Luxburg UV, Bengio S, Wallach H, Fergus R, Vishwanathan S, et al., editors. Advances in Neural Information Processing Systems 30. Curran Associates, Inc.; 2017. p. 4765–74.

31. Hähnel T, Baldow C, Guilhot J, Guilhot F, Saussele S, Mustjoki S, et al. Model-Based Inference and Classification of Immunologic Control Mechanisms from TKI Cessation and Dose Reduction in Patients with CML. Cancer Res. 2020. p. 2394–406.

32. Chollet F. keras [Internet]. Github; [cited 2020 Nov 10]. Available from: https://github.com/keras-team/keras

33. Chaloner K, Verdinelli I. Bayesian Experimental Design: A Review. Stat Sci. Institute of Mathematical Statistics; 1995;10:273–304.

34. Goodwin. Dynamic System Identification: Experiment Design and Data Analysis. Academic Press; 1977.

35. Seeger MW. Bayesian Inference and Optimal Design for the Sparse Linear Model. J Mach Learn Res. 2008;9:759–813.

36. Walter E, Pronzato L. Identification of Parametric Models: from Experimental Data. Springer London; 2010.

37. Clark RE, Polydoros F, Apperley JF, Milojkovic D, Rothwell K, Pocock C, et al. De-escalation of tyrosine kinase inhibitor therapy before complete treatment discontinuation in patients with chronic myeloid leukaemia (DESTINY): a non-randomised, phase 2 trial. Lancet Haematol. 2019;6:e375–83.

38. Chen RTQ, Rubanova Y, Bettencourt J, Duvenaud D. Neural Ordinary Differential Equations [Internet]. arXiv [cs.LG]. 2018. Available from: http://arxiv.org/abs/1806.07366

39. De Brouwer E, Simm J, Arany A, Moreau Y. GRU-ODE-Bayes: Continuous modeling of sporadically-observed time series. arXiv [csLG] [Internet]. 2019; Available from: https://arxiv.org/abs/1905.12374

40. Arras L, Montavon G, Müller K-R, Samek W. Explaining Recurrent Neural Network Predictions in Sentiment Analysis [Internet]. arXiv [cs.CL]. 2017. Available from: http://arxiv.org/abs/1706.07206

41. Shrikumar A, Greenside P, Kundaje A. Learning Important Features Through Propagating Activation Differences [Internet]. arXiv [cs.CV]. 2017. Available from: http://arxiv.org/abs/1704.02685

42. Esteva A, Robicquet A, Ramsundar B, Kuleshov V, DePristo M, Chou K, et al. A guide to deep learning in healthcare. Nat Med. 2019;25:24–9.

43. Rudin C. Stop explaining black box machine learning models for high stakes decisions and use interpretable models instead. Nature Machine Intelligence. 2019;1:206–15.

